# Sports and exercise medicine clinic in public hospital settings: A real-life concept and experiences of the treatment of the first 1151 patients

**DOI:** 10.1101/2022.06.13.22276140

**Authors:** Lauri Alanko, Jari A. Laukkanen, Mirva Rottensteiner, Salla Rasmus, Tero Kuha, Maarit Valtonen, Urho M. Kujala

**Affiliations:** Central Finland Health Care District, Sports and Exercise Medicine Clinic, Jyväskylä, Finland; Faculty of Sport and Health Sciences, University of Jyväskylä, Jyväskylä, Finland; Finnish Institute of High Performance Sport KIHU, Jyväskylä, Finland; Institute of Clinical Medicine, Department of Medicine, University of Eastern Finland, Kuopio, Finland; Central Finland Health Care District, Department of Medicine, Jyväskylä, Finland

**Author notes:** Correspondence to: Urho M. Kujala, Faculty of Sport and Health Sciences, University of Jyväskylä, P.O. Box 35, FI-40014 Jyväskylä, Finland.

## Abstract

Exercise has been shown to have a multitude of health-promoting effects, including improvements in cardiorespiratory and muscular fitness, function, symptoms and risk factors. Furthermore, exercise may slow down, stop or even reverse the progression of various non-communicable diseases (NCDs). Despite the overwhelming evidence, exercise is still not comprehensively used as a treatment component either in primary care or in hospital settings. The outpatient Sports and Exercise Medicine Clinic (SEMC) is the first specialized clinic in Finland to use exercise as part of patient protocols in the public health care system. Patients needing specialist attention due to NCDs, usually combined with sedentary lifestyles, are referred to this central hospital clinic. The prerequisites for patient referral are the known efficacy of exercise intervention in the treatment of disease and the need for sports and exercise medicine expertise. The focus of the clinic is to implement regular physical activity into daily life and give guidance on other health-promoting habits such as diet, rest and the reduction of substance use. In addition, SEMC promotes the inclusion of exercise in several local treatment guidelines in the hospital district, such as the treatment paths of non-insulin-dependent diabetes, sleep apnoea, severe obesity indicating bariatric operation, and breast cancer. The advisory treatment protocol of SEMC consists of a primary evaluation, face-to-face controls with a physician and/or physiotherapist at 3, 6 and 9–12 months, and contacts via phone between hospital visits. Laboratory tests, body composition, walking tests, and measurements of muscle strength and balance are performed at the beginning of the treatment. Body composition and physical tests are repeated after 6 and 9–12 months. In addition, questionnaires on lifestyle and quality of life are completed prior to the first visit and at 6 months of control. At the core of the treatment is individualization, using motivational interviewing, considering the patients’ personal interests and possibilities, and encouraging the patient to be an active member of our multi-professional team. In this article, our protocol is described and some preliminary data and experiences on the treatment of first 1151 patients are reported. Furthermore, strengths, challenges and future steps in the development of our protocol and outpatient clinic are discussed.

**Key messages box:** Randomized controlled trials show that exercise therapy improves function, the cardiometabolic risk factor profile, mood, some indicators of disease progression, and health-related quality of life in patients with a variety of chronic diseases.

However, exercise therapy has only occasionally been incorporated into the treatment of chronic diseases in public healthcare organizations.

We summarize experiences of the clinic development and treatment of patients at the Sports and Exercise Medicine Clinic at the Central Hospital of Central Finland.

Our protocol consists of a primary evaluation, face-to-face controls with a physician and/or physiotherapist at 3, 6 and 9–12 months, contacts via phone between hospital visits, as well as the use of questionnaires, body composition, and fitness tests during the contacts.

At the core of the treatment are personalized exercise recommendations that are tailored using motivational interviewing and taking the patient’s personal interests, possibilities and limitations into consideration.

Collaboration with primary care and exercise-sector professionals in the hospital district makes it possible to tailor exercise therapy to many patients for both prevention measures and the treatment of the patients.

## Background

A sedentary lifestyle has been shown to increase the prevalence of several chronic diseases and risk factor levels, such as type 2 diabetes, elevated blood pressure, obesity and sleep apnoea. Yet, there is strong evidence from randomized controlled trials (RCTs) that exercise therapy leads to improved physical fitness and function in patients with chronic diseases (Pasanen et al., 2017) and has other health benefits as well (Kujala, 2009; Pedersen and Saltin, 2015). Exercise training leads to improvements in body composition, including body weight control and the reduction of visceral fat (Ohkawara, 2007; Kim et al., 2019). There are meta-analyses of RCTs in patients with NCDs summarizing the evidence of other multi-dimensional benefits of exercise therapy (Kujala, 2021). These meta-analyses show that various cardiometabolic risk factor levels are improved in most of the common cardiometabolic diseases, pain is reduced in musculoskeletal diseases, mood (depression and anxiety) and health-related quality of life are improved in various disease categories, and disease-specific ‘proxy’ indicators of disease progression are improved in diseases such as type 2 diabetes, hypertension, coronary heart disease, heart failure, claudication, chronic obstructive pulmonary disease, rheumatoid arthritis, fibromyalgia, depression, anxiety and schizophrenia (Kujala, 2021).

Despite the overwhelming evidence, the systematic implementation of exercise therapy in medical care has been limited. Healthcare professionals may have been reluctant to recognize exercise as a part of medical treatment. Systematic use of exercise training has been thought to belong more to the world of sports, and its wider implementation from younger to adult populations is thought to be the mission/duty of organizations other than the healthcare system. There are different types of sports and exercise medicine clinics, but usually they are not linked to public health care, which would give all patients with NCDs the opportunity to access exercise therapy, and clinics for specific diseases have only randomly included exercise in their treatment protocols. This paper describes the protocol and preliminary results of the recently developed Sports and Exercise Medicine Clinic (SEMC) in the Central Finland Central Hospital (CFCH), discusses experiences and offers suggestions for further development.

## Methodological development and concept of the sports and exercise medicine clinic

### Patient scope and referral

The SEMC is open to all patients in the Central Finland Health Care District (total population: 252,676), and they are referred to SEMC by both primary health care and CFCH physicians and also from private health care services. The referral criteria are the evidence-based effect of exercise in the treatment of patients’ disease, the need for specialist-led exercise interventions for patients with exercise disabling NCDs, and patients’ own even mild to moderate motivation to implement exercise into their everyday lives. A typical patient has several NCDs and a sedentary lifestyle.

### Steps in the clinic development

From the beginning, SEMC has been collaborating actively with the Faculty of Sports and Health Sciences of the University of Jyväskylä. The first step in the development of the SEMC in the CFCH was a questionnaire study offered to all specialist physicians at the hospital. All respondents supported the development of the clinic and indicated that physical inactivity with obesity was the most common challenge needing attention (Backman, 2005).

After piloting the project, the clinic was started in 2016 with a part-time specialist physician in Sports and Exercise Medicine (SEM) and a part-time physician specializing in SEM; later, a part-time physiotherapist was added. Currently, the personnel consist of a part-time chief physician (specialist in SEM), a full-time physician in training and a full-time physiotherapist. Throughout the years, the protocol has been tested and modified in order to improve it.

The ethics committee of the Central Finland Health Care District, Jyväskylä, Finland, has evaluated the reporting of the treatment results of the SEMC patients and gave permission to the study through commenting that there is no need for a specific ethics committee statement as the study reports results of the hospital’s accepted normal treatment praxis and only the hospital physician has access to patient data with patient identification codes. Then the Central Finland Central Hospital gave permission to this SEMC study.

### Current concept of the SEMC

The exercise intervention protocol usually lasts 9–12 months and consists of a primary visit and follow-ups at 3, 6 and 9–12 (the last one is decided individually) months after the primary visit. In between visits to the SEMC, patients are contacted by phone.

The primary visit lasts 60 minutes, during which the patient’s overall situation is evaluated. In addition to possible routine disease-specific medical examinations, prior to the first and 6-month visits to the clinic, patients complete both the lifestyle questionnaire (including questions on exercise, sitting time, dietary habits, sleep, alcohol consumption, smoking) and the quality of life questionnaire RAND-36 (Hayes, 1993). The medical conditions are evaluated, as well as the possibilities for increasing exercise and developing a healthier lifestyle. Laboratory tests (including an ECG) are collected prior to the first visit to assist in the medical evaluation of the patient’s risk factors. By the end of the visit, goals are set and recorded. The goals, which are tailored to each patient individually, include the amount of increase in activity/exercise, possible changes in diet and decrease in substance use, screen time, and so forth. The forms of activity are decided based on the interests of the patient, considering possible medical/economic/social hindrances. The patient helps set these goals and is thus included in the treatment decision-making. The principles of motivational interviewing are applied throughout the process.

The American College of Sport Medicine’s (ACSM) recommendation for exercise is 2.5 hours of aerobic training and two strength training sessions a week (Physical Activity Guidelines Resources (acsm.org)). At the start of the exercise intervention this recommendation is often unrealistic for SEMC patients, as most of them have previously engaged in little or no physical activity. In general, the main goal at the beginning of the intervention is getting any exercise at all to become a regular part of a patient’s everyday life with later progression of the exercise amount.

The physician’s appointment is followed by two hours of detailed assessment and the evaluation of functional capacity and body composition by an experienced physiotherapist. The tests include a 6-minute walking test (occasionally, a stress-ECG test is done prior to a walking test for patient safety), body composition analysis (bioimpedance, Biospace InBody 770), sit-ups from a chair (with patients over 55 years), squats (with patients 35–55 years), hand-grip strength and one-leg stance. In the case of a specific need, either a medical or psychological or another appointment with the physiotherapist is scheduled. These visits are usually either to treat a musculoskeletal disease (i.e. osteoarthritis, chronic lower back pain, etc.) or to develop a personal exercise and/or strength programme, especially in cases when attending a gym is not possible (due to distance, financial or social issues, etc.).

To further help the patient in the beginning, SEMC has its own strength-training group. This group is a very low-level entry for those who have no previous experience in weight training and who usually find it very hard to go to an open gym. There are six patients in this group at a time, enabling personal supervision by our physiotherapist. Patients can take part in this group six times, after which they are guided to communal and/or private gyms that suit the patient in question. In addition, SEMC collaborates with municipal recreation services, taking into account the restrictions in exercising. In the communal services, patients receive personal assistance when starting to exercise, information about public exercise facilities/opportunities and even follow-up visits.

During control visits, patients’ progress is evaluated. Positive feedback is given even for patients with minor advances and possible problems are assessed. The goals are re-evaluated and, if necessary, re-adjusted. At 6 months, the effects of intervention are re-evaluated, and if it is obvious that no progress is forthcoming or that the goals of the intervention have been achieved, the intervention is terminated. It is rare that after 6 months, the patient has both implemented so much exercise into his/her everyday life that the ACSM guidelines have been fulfilled and the patient is mentally ready to continue the physically more active lifestyle on her/his own. However, usually, the follow-up ends after 9–12 months, and even if the positive changes are excellent after 6 months, the patient still needs psychological support. Sometimes the progress begins to happen later in the intervention, and in these cases, the intervention can be prolonged. The decision to end the contacts at SEMC is again made together with the patient, not solely by the physician.

Additionally, there are nutritional therapists both in the hospital and in the primary health care, and patients with a BMI over 35 can be directed for consultation.

The latest addition to the protocol is a joint project with LIKES Research Centre on Physical Activity and Health, in which patients with the lowest scores in the RAND36 survey have the opportunity to meet with a certified psychologist.

## Experiences and comments

Since the start of autumn 2016 to end-December 2021, the SECM has treated 1151 patients (Figure 1). The patient flow rose in the first years and has plateaued since then, reaching the current capacity limit. Table 1 lists the 20 most common diagnoses of our 2016–2020 patients. Overweight is present in approximately two-thirds of all patients, followed by (co-existing) sleep apnoea, hypertension, non-insulin dependent diabetes, and asthma. The first musculoskeletal disorder in which symptoms are worsened by physical inactivity and attenuated with exercise – osteoarthritis – is ranked sixth. Cardiovascular disease-related diagnoses are less common than expected, as a separate physiotherapy-led rehabilitation protocol for cardiac patients already exists in the hospital district.

**Table 1.** Typical patient groups which have been forwarded to the Sports and Exercise Medicine clinic, with the main goals of treatment and simple follow-up parameters in use or recommended.

| **Disease** | **Main goals of the exercise therapy** | **Easy-to-use follow-up measures\*** | **N†** |
| --- | --- | --- | --- |
| Obesity, including overweight in children | Reduction of/controlling body weight, prevention of metabolic disturbances, maintaining muscle mass during weight reduction | WT, WM, body composition, muscular strength tests, glucose and lipid risk factors | 1011 |
| Sleep apnoea | Reduction of daytime tiredness, improving sleep quality, reduction of weight, improving fitness | WT, WM | 637 |
| Hypertension | Reduction of blood pressure levels and weight, treatment of metabolic disturbances | Blood pressure, body composition | 384 |
| Type 2 diabetes | Improving glucose metabolism, preventing complications | WM, glucose and lipid levels | 228 |
| Asthma | Improvement in exercise capacity and quality of life, reduction of dyspnoea symptoms | WT, clinical exercise test, symptom questionnaire, PEF follow-up, flow-volume spirometry | 186 |
| Arthrosis and other degenerative joint diseases | Pain reduction and function improvement | Pain-VAS, WT, muscular strength tests | 127 |
| Metabolic syndrome | Improvements in metabolism, reduction of waist circumference, improvement in cardio-metabolic risk factors and physical fitness | WT, WM, blood glucose and lipids, blood pressure | 120 |
| Chronic pain in any part of the spine, with or without neural compression, spondylolisthesis, stenosis etc | Pain reduction, function improvement | Pain-VAS, muscular strength tests | 103 |
| Hypercholesterolemia | Improvement in lipid profile | blood lipids | 94 |
| Neurologic diseases (MS, Parkinson's disease, etc.) | Maintaining or improving mobility, motor control and balance | WT, balance tests | 84 |
| Coronary heart disease | Prevention of disease progression, reduction of symptoms | WT, WM, glucose and lipid risk factors | 81 |
| Hypothyroidism | Reduction of symptoms | WT, blood test (TSH, T4v) | 80 |
| Atrial fibrillation | Control of symptoms, maintaining exercise- and functional capacity | WT, heart rate response during exercise test | 76 |
| Depression | Reduction of depression symptoms | Mood questionnaire | 48 |
| Tendinopathies, bursitis, tendon overuse injuries | Pain reduction and function improvement | Pain-VAS, WT, muscular strength tests | 38 |
| Fibromyalgia | Maintaining physical fitness, mood, and function | WT, symptom questionnaire | 32 |
| Acute myocardial infarction, rehabilitation | Improving physical fitness and functional capacity<br>Reducing symptoms/symptom control and possible fears of physical activity | WT, WM, heart rate responses during exercise and assessment of CVD risk factors | 31 |
| Cancer | Maintaining or improving physical and mental function, muscle mass and quality of life | WT, muscular strength tests | 26 |
| Chronic heart failure | Symptom control, improvement or maintenance of function, prevention of disease progression | WT, heart rate response during exercise, blood tests (pro-BNP) | 24 |
| Bi-polar disorder | Reduction of symptoms | Mood questionnaire | 21 |

\*Disease-specific quality of life questionnaires are important in the evaluation of treatment results.
†Diagnosis number among 1,535 first patients visiting the clinic (one patient can have several diagnoses)
WT=walking test, WM=measurement of waist circumference, VAS=visual analogue scale.

**Figure 1.**
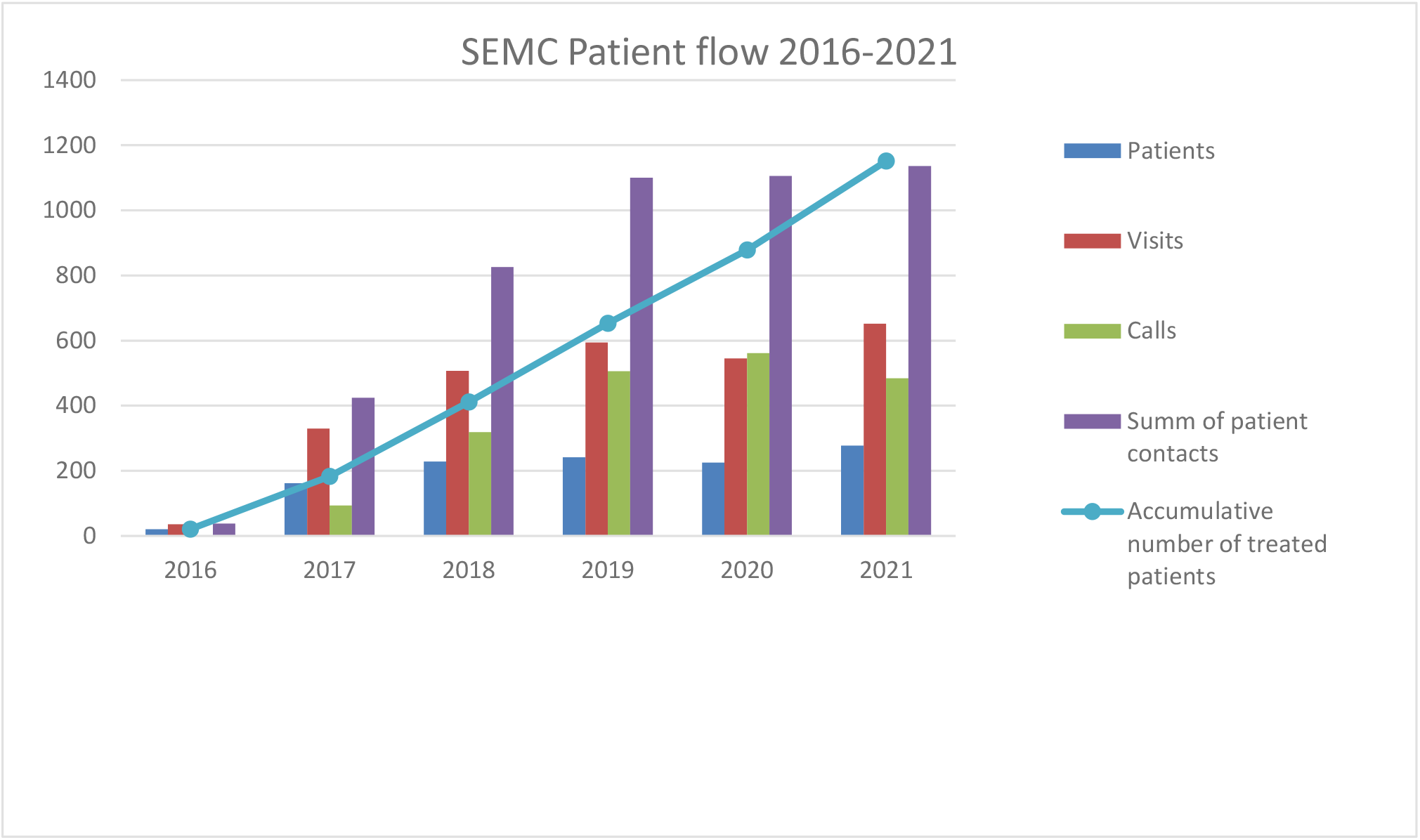
The number of yearly new patient visits (blue), all yearly patient visits (maroon), yearly phone calls (green), all yearly patient contacts (violet), and cumulative number of treated patients (light blue line) from 2016 through 2021.

In its first five years (from 2016 to 2020) SEMC treated 874 patients, with 457 different diagnoses adding to the total number of 4,158 diagnoses, that is, 4.8 diagnoses per patient. Thus, the SEMC patient sample deviates from typical patients in exercise therapy RCTs, who commonly have only one disease.

Personalized exercise therapy has been the core protocol, adjusted to patients’ needs, abilities and opportunities. Patients have given positive feedback, and quite a few have succeeded in changing their sedentary lifestyle into a physically active one. A rough estimate based on three master’s theses produced by SEMC patients Renkola (2020), Laaksonen (2020) and Ben-Khalifa (2021) and the personal views of the personnel working in the SEMC is that slightly more than half of the patients succeeded in increasing exercise clinically significantly. Thus, while our real-life experience indicates that this is somewhat lower than what has been reported in RCTs, the patients at SEMC have not been excluded on virtually any criteria such as low motivation, and the volume, intensity and type of exercise has been tailored to severely incapacitated patients as well.

Individuals have different interests regarding types of exercises, and our experience is that it is critical to discuss the likes/dislikes of the patient, ask for preferences and be supportive in the choice of activities. The most common activity is walking/Nordic walking, followed by swimming/water running, gym training, (electric-)cycling and cross-country skiing. But there are patients who participate in dancing and even virtual reality gaming. In short, if a patient likes an activity that is not a health risk, he/she should be encouraged. The starting activity levels vary from none to several hours of weekly exercise to several hours but are usually below the national guidelines. Another typical feature in SEMC patients is that there can be some aerobic exercise or strength training at the beginning of the intervention, but very rarely both. The level of effort in aerobic training is low, that is, slow walking with no/little effort. One of the goals during the intervention is to guide the patient into the existing exercise facilities, non-supervised and supervised, instead of organizing separate training programmes. In this way, we try to reach a high cost-benefit ratio and promote long-term lifestyle change.

Furthermore, according to our experiences at SEMC, there are patients who start increasing physical activity during their waiting time for the clinic or are very responsive to the physician’s simple advice. These patients have often been physically active during childhood/adolescence but changed later to sedentary lifestyles. On the other end, there are patients who have never been physically active in their entire lives and need more support both psychologically and in starting physical exercise. At the core of personalized exercise therapy is that the provision of therapy can and should be intensified or changed during follow-up appointments if the patient has not been able to implement exercise into his/her everyday life. In our experience, this kind of treatment protocol may lead to better overall long-term success than that reported in RCTs with strictly standardized programmes, especially with patients with several NCDs and disabilities.

So far, SEMC relies on the activity level reported by the patient (through interviews and questionnaires). Most SEMC patients do not have sport watches or activity trackers. Some patients find them a positive challenge, while others consider them more of a threat, causing psychological pressure. On the other hand, an old-fashioned exercise diary has proven to be an easy and cost-effective way of monitoring and encouraging daily activities. Different types of technology are needed to meet individual preferences.

RCTs are the most common method of evaluating the effects of treatment, but they have some serious downsides from the real-life perspective. First, participants for RCTs are usually selected and highly motivated, whereas those with the gravest need for increased physical activity are probably the least likely to participate RCTs as some of the SEMC patients with increased age and various NCDs. Reasons for being physically inactive are multitude including such as having several NCDs, social/financial difficulties. Second, in reported RCTs, all intervention group participants have usually followed the same pre-determined intervention, thus neglecting the personal interests and what may or may not be possible for the patient – given the circumstances/health/age and so on. In our experience, the physician/physiotherapist must first agree with the patient on personalized ways of increasing physical activity to motivate the individual. Identifying the type of intervention according to personal motives, hopes, needs and possibilities for increasing physical activity may lead to a better success rate, cost effectiveness and results in real life compared to a strict one-programme-fits-all principle.

More critical analyses of the effectiveness of SEMC treatment protocol are to come, but we have monitored the effectiveness of the treatment to get feedback for further development. Among other things, patients have been cured of sleep apnoea, have had their type 2 diabetes put into remission, have had their anxiety disorder/depression attenuated, and have increased their aerobic capacity, balance and strength, with or without weight loss. At the beginning of the SEMC development, there was close collaboration with the clinic for pulmonary diseases, and follow-ups show that sleep apnoea patients benefit from exercise therapy, also independent of weight reduction (Renkola, 2020), which is in line with recent RCTs. As expected, reduction of body weight and fat percentage are associated with the increases in physical activity levels of the patients (Laaksonen, 2020), but it should be noted that the health benefits of regular exercising also occur independent of weight reduction in most patient groups.

### Integration of exercise into usual treatment paths

Due to the wide range of effects of exercise on several non-communicable diseases, SEMC has been actively involved in the development of several local treatment guidelines for CFCH. In the treatment path for sleep apnoea patients with several NCDs, for whom the CPAP treatment is not successful/patient does not want CPAP treatment or when the overall situation requires specialist-led intervention, they are offered the opportunity to receive an SEMC exercise intervention protocol. In the type 2 diabetes treatment path, the SEMC exercise intervention is offered to patients with comorbid disease that is hindering exercise. All patients with morbid obesity indicating bariatric surgery are directed to a slightly modified exercise intervention. In addition, all mammary cancer patients with post-operative hormonal treatments are offered the opportunity of an SEMC consultation, a single visit, a full protocol, or a personally adjusted one. SEMC is collaborating with primary health care to develop treatment guidelines and protocols to include exercise as part of the treatment of all patients with insufficient exercise levels.

## Future considerations

The SEMC protocol has been developed and modified throughout its existence, with the ultimate goal of becoming effective, cost-effective and monitorable and able to be implemented in other health care districts in Finland (and even worldwide). Physical capacity tests and body composition analyses are validated methods, easy to perform and repeatable. The addition of questionnaires to the protocol made it easier to follow changes in exercise, physical activity, diet and subjective quality of life. Currently, we are working to standardize the way patient reports are written. We are planning to collect automated feedback from the patients to gain more insight into which parts of the protocol the patients consider most valuable and what changes they would prefer.

SEMC is collaborating with the University of Jyväskylä in doing research on the effectiveness of the SEMC protocol. As mentioned above, so far, three master’s theses have been written by patients of SEMC. To create a sound platform for future research and a tool to develop the protocol, a detailed real-life database that includes all our patients is under development. This database will be used both to assess the efficacy of the treatment and to evaluate the protocol, thus leading to development in local clinical practices.

In the long run, we would like to increase the SEMC multi-professional team to include new specialists, above all, a full-time psychologist. The responses from the project with the LIKES Research Centre on Physical Activity and Health have been positive, and there would be a great demand for psychological support. In addition to patient work, the psychologist would be a much-needed support for the other staff members to increase their skills in supporting the patient. Another much-needed specialist is a nutritional therapist; the need at the hospital exceeds capacity at the moment. Here, too, the therapist could be consulted by the rest of the team more easily than is now possible. A physiologist would be needed to take at least part of the measurements, to conduct testing with the patients and to develop the methods. This would free up time for the physiotherapist. Something that is very much needed and hoped for by the patients is more strength training groups, which would require more physiologist/physiotherapy capacity. An easy transition from the SEMC training group to public gyms and exercise facilities still needs an improved protocol, a matter to be worked on with the communal exercise professionals. Additionally, it is vital that continuous monitoring of the process and results, combined with a constant drive to evolve SEMC, is further developed. This will be done using upcoming patient register studies in collaboration with the Faculty of Sports and Health Sciences, University of Jyväskylä.

We will also continue to collaborate with primary health care personnel in Central Finland to actively screen for patient’s physical activity levels and take action by delivering personalized exercise therapy if the individual’s physical activity does not meet the national recommendations. So far, two primary care physicians have had their three-month training periods in SEMC. In two primary health care centres, there are exercise instructors working within the health care system, and more will follow. An important upcoming step in the development of collaboration has been the addition of an exercise report to the electronic patient record in which both health care and exercise professionals can document and follow the progress of the exercise therapy.

## Conclusions

Personalized exercise therapy is not being used as a medical treatment to its full potential. Developing and implementing personalized exercise therapy as part of the healthcare system is of major interest, and it may have a worldwide impact, be cost-efficient for the community and effectively improve an individual’s health and quality of life.

The SEMC protocol is based on individual exercise implementation, developed by both the patient and a multi-professional team. In the SEMC experience, it is possible to increase a patient’s physical activity level, subjective wellbeing and quality of life, even if the patient has no history of physical activity and has a multitude of disabling NCDs. We have gained encouraging experience as we seek to realize the goal of integrating and linking personalized exercise closely to patient treatment in both hospital and primary health care settings.

## Data Availability

All data produced in the present study are available upon reasonable request to the authors

## References

1. Pasanen T, Tolvanen S, Heinonen A, Kujala UM. Exercise therapy for physical function in chronic diseases: a systematic review of meta-analyses of randomized controlled trials. Br J Sports Med 2017;51:1459–65.

2. Kujala UM. Evidence of the effects of exercise therapy in the treatment of chronic disease. Br J Sports Med 2009;43:550–55.

3. Pedersen BK, Saltin B. Exercise as medicine – evidence for prescribing exercise as therapy in 26 different chronic diseases. Scand J Med Sci Sports 2015;25 (Suppl. 3):1–72.

4. Ohkawara K, Tanaka S, Miyachi M, Ishikawa-Takata K, Tabata I. A dose-response relation between aerobic exercise and visceral fat reduction: systematic review of clinical trials. Int J Obes (Lond) 2007;31:1786–97.

5. Kim KB, Kim K, Kim C, Kang SJ, Kim HJ, Yoon S, et al. Effects of exercise on the body composition and lipid profile of individuals with obesity: A systematic review and meta-analysis. J Obes Metab Syndr. 2019;28:278–94.

6. Kujala UM. Summary of the effects of exercise therapy in non-communicable diseases: Clinically relevant evidence from meta-analyses of randomized controlled trials. MedRxiv 2021; doi: https://doi.org/10.1101/2021.02.11.21251608.

7. Backman K. Physicians’ views on the need of an exercise clinic based in the Central Finland Central Hospital (In Finnish with an abstract in English). Faculty of Sport and Health Sciences, University of Jyväskylä, Master’s thesis, 2005. http://urn.fi/URN:NBN:fi:jyu-200540

8. Hayes RD, Sherbourne CD, Mazel RM. The RAND 36-item Health Survey 1.0. Health Economics 1993;2:217–27.

9. Renkola H-K. The impact of individualized exercise program on overweight sleep apnea patient’s symptoms, anthropometry and functional capacity: Results of the sports and exercise medicine outpatient clinic (In Finnish with an abstract in English). Faculty of Sport and Health Sciences, University of Jyväskylä, Master’s thesis, 2020. http://urn.fi/URN:NBN:fi:jyu-202004272902.

10. Laaksonen M. Effects of exercise treatment on anthropometry, physical functioning and perceived health among obese patients in the Sport and Exercise Medicine outpatient clinic (In Finnish with an abstract in English). Faculty of Sport and Health Sciences, University of Jyväskylä, Master’s thesis, 2020. http://urn.fi/URN:NBN:fi:jyu-202007165342.

11. Ben-Khalifa J. Effect of exercise therapy on physical function, body composition and increased physical activity directed at the Sports and Exercise Medicine Clinic (In Finnish with an abstract in English). Faculty of Sport and Health Sciences, University of Jyväskylä, Master’s thesis, 2021. https://jyx.jyu.fi/handle/123456789/76354.

